# A quantitative risk-benefit analysis of ChAdOx1 nCoV-19 vaccine among people under 60 in Italy

**DOI:** 10.1101/2021.05.07.21256826

**Authors:** Raffaele Palladino, Daniele Ceriotti, Damiano De Ambrosi, Marta De Vito, Marco Farsoni, Giuseppina Seminara, Francesco Barone-Adesi

## Abstract

ChAdOx1 nCoV-19 is a vaccine against the COVID-19 infection that was granted a conditional marketing authorization by the European Commission in January 2021. However, following a report from the Pharmacovigilance Risk Assessment Committee (PRAC) of European Medicines Agency, which reported an association with thrombo-embolic events (TEE), in particular Disseminated intravascular coagulation (DIC) and Cerebral venous sinus thrombosis (CVST), many European countries either limited to individuals older than 55-60 years or suspended its use. We used publicly available data to carry out a quantitative risk-benefit analysis of the vaccine among people under 60 in Italy. Specifically, we used data from PRAC, Eudravigilance and ECDC to estimate the excess number of deaths for TEE, DIC and CVST expected in vaccine users, stratified by age groups. We then used data from the National Institute of Health to calculate age-specific COVID-19 mortality rates in Italy. Preventable deaths were calculated assuming a 72% vaccine efficacy over an 8-month period. Finally, benefit-risk ratio of ChAdOx1 nCoV-19 vaccination was calculated as the ratio between preventable COVID-19 deaths and vaccine-related deaths, using Monte-Carlo simulations. We found that among subjects aged 20-29 years the benefit-risk [B-R] ratio was not clearly favorable (0.70; 95% Uncertainty Interval [UI]: 0.27-2.11). However, in the other age groups the benefits of vaccination largely exceeded the risks (for age 30-49, B-R ratio: 22.9: 95%UI: 10.1-186.4). For age 50-59, B-R ratio: 1577.1: 95%UI: 1176.9-2121.5). Although many countries have limited the use of the ChAdOx1 nCoV-19 vaccine, the benefits of using this vaccine clearly outweigh the risks in people older than 30 years. The use of this vaccine should be a strategic and fundamental part of the immunization campaign considering its safety and efficacy in preventing COVID-19 and its complications.

## Introduction

The Oxford-Astrazeneca ChAdOx1 nCov-19 vaccine is a recombinant chimpanzee adenoviral vector encoding the SARS-CoV-2 spike glycoprotein [1]. Trial evidence suggests a 72% efficacy in preventing the COVID-19 infection [2]. Vaccine marketing authorization was provided by the European Medicine Agency (EMA) on January 29, 2021 [3], while the World Health Organization (WHO) approved its use on February 15, 2021 after it was evaluated by WHO’s Strategic Advisory Group of Experts on Immunization (SAGE) [4]. Following this evaluation, this two-dose vaccination was initially recommended for individuals over the age of 18 [5]. Majority of the European countries decided to administer it mainly for those under the age of 60 with some small differences between countries [3].

In March 2021, the use of ChAdOx1 nCoV-19 vaccine was paused in a number of European countries due to the reports of thromboembolic events (TEE), in particular involving unusual locations such as cerebral venous sinus and splanchnic circulation [6]. According to the first reports, these rare events tend to appear within two weeks after administration of the vaccine [3].

Even if EMA has not changed since then its indications on the recommended age groups to receive the vaccine [3], many countries, including Italy, currently recommend the use of vaccination only for subjects over 60 years, without prejudice to the need to give the second dose to younger subjects who have already received the first and have not presented adverse events. This decision is having obvious negative consequences for the immunization campaign, already slowed down by problems with the supply of all the approved vaccines, especially in those European countries which have been impacted the most by the pandemic.

EMA has recently conducted a preliminary risk-benefit analysis of the ChAdOx1 nCov-19 vaccine in the adult population using data from European Union Member countries to compare the risk of TTE with the expected vaccination benefits in preventing COVID-19 and its complications [7]. Results suggests that benefits outweigh risks in individual aged at least 40 years, but not in younger people. However, we argue that the benefit-risk ratio might be even more favorable for those countries, such as Italy, characterized by a large circulation of the virus and high case fatality rate. For this reason, we used real-world data from Italy to carry out a comprehensive, country specific, benefit-risk analysis ChAdOx1 nCov-19 vaccine in different age groups.

## Methods

### Parameters of the simulation

The benefit-risk analysis of the vaccination was carried using Monte Carlo simulations, based on different parameters that we estimated from publicly available data and that are described hereafter (table1):

#### Vaccine efficacy

we assumed a 72% efficacy of the vaccination, as reported by Emary and colleagues [2]. As studies suggest that Covid-19 vaccine-induced immunity is at least six months [8], in our calculation we conservatively assumed vaccination being effective for eight months.

#### Covid-19 mortality rates

we used weekly reports of the Italian National Health Institute to obtain the number of Covid-19 deaths occurred during the last five months (from 25/11/2020 to 13/04/2021) [9-10]. The age structure of the Italian population was obtained by the database of the National Institute of Statistics (ISTAT) [11]. Age-specific mortality rates and their 95% confidence intervals were estimated assuming a Poisson distribution of the events.

#### Baseline Incidence rates of thrombo-embolic events (TEE), disseminated intravascular coagulation (DIC) and cerebral venous sinus thrombosis (CVST)

Age-specific rates in the general population were retrieved by the PRAC report and associated documentation [3-12].

#### Relative risks of TEE/DIC/CVST in vaccine users

age-specific relative risks of different adverse thrombotic events during the 14 days following the vaccination were retrieved by the PRAC report [3].

#### TEE/DIC/CVST fatality rates

fatality rates (and their 95% CI) of the different adverse thrombotic events were computed using the number of deaths over the total number of cases reported in the PRAC report and assuming a binomial distribution of events [3].

### Updated values of the parameters

As the data reported in the PRAC report included only adverse events occurred among ChAdOx1 nCov-19 vaccine users until March 16th, we conducted a new search in the Eudravigilance database [13] on April 28^th^. This allowed to increase the number of identified cases of DIC and CVST from 7 and 18 to 27 and 116, respectively, and obtain more precise estimates of vaccine-associated relative risks and fatality rates for these conditions. Data on DIC were obtained searching in the section “Blood and lymphatic system disorders’’ of reported suspected reactions associated with the words “disseminated intravascular coagulation”, while CVST data were obtained searching suspected reactions in the section “Nervous system disorders’’ associated with the words “cerebral venous sinus thrombosis”. Data on the number of vaccinated subjects in the EEA countries (necessary to estimate the incidence rates of adverse events among vaccine users) were obtained from the European (ECDC) database [14]. For some countries (Cyprus, Germany, France, Liechtenstein, the Netherlands, Norway, Romania, and Slovakia), only the overall data of vaccinated subjects, without subdivision by age group, were available. In this case, the number of subjects in the different age-groups was calculated using the age-distribution of vaccinated reported in the other EEA countries. Also, the cut-offs of age-groups used in the ECDC website were slightly different from those used in the PRAC report. For this reason, we applied the 20-29 and 30-49-years baseline rates of DIC and CVST to the rates of the 18-24 years and 25-49 years age-groups of vaccinated subjects to estimate the corresponding relative risks.

The benefit-risk ratio of ChAdOx1 nCoV-19 vaccination was calculated as the ratio between preventable COVID-19 deaths and vaccine-related deaths. We compared the results considering all TEE events and those restricted only to DIC and CVST (these two conditions being apparently more strongly associated to ChAdOx1 nCov-19 vaccine [3]). The analysis restricted to DIC and CVST was also re-run using the updated values of the parameters. Finally, we conducted secondary analysis assuming different degrees of under-reporting of TEE.

To evaluate the uncertainty of our estimates, in each simulation we sampled the values of the parameters from their confidence intervals. We generated 95% uncertainty intervals (95%UI) by taking the 2.5% and 97.5% percentile estimates from 100,000 simulations. We also plotted the expected benefits and risks of each simulation to better evaluate the safety profile of the vaccine (probabilistic sensitivity analysis). All statistical analyses were performed using Stata version 15 (StataCorp, College Station, TX).

### Ethics Committee Approval

The study was based on publicly available aggregate data. No ethics committee approval was necessary.

## Results

The results of the benefit-risk assessment of ChAdOx1 nCoV-19 vaccination are shown in Table 2. Using data based on the PRAC report, we found that among subjects aged 20-29 years the benefit-risk ratio was not favorable, neither considering TEE (0.70; 95%UI 0.27-2.11), nor DIC-CVST (0.93; 95%UI 0.06-14.3). However, in the other age groups the benefits of vaccination largely exceeded the risks. In particular, in the 30-49 age group, the estimated benefit-risk ratio for TEE was 22.9 (95%UI:10.1-186.4) while for DIC and CVST was 52.2 (95%UI:16.6-179.1). In the 50-59 age group the benefit-risk ratio was even larger (for TEE 1577.1; 95%UI:1176.9-2121.5; for DIC and CVST 3506.7; 95UI:166.0-54427.9). These results were confirmed in the analysis based on the updated data, that clearly showed a benefit for vaccine users over age 24 (table 2).

**Table 1.** Parameters used for Monte Carlo simulations.

| Parameter | Original estimates |  | Updated estimates |  |
| --- | --- | --- | --- | --- |
|  | Estimate (95%CI) | Ref. | Estimate (95%CI) | Ref. |
| Vaccine Efficacy | 0.72 (0.63-0.79) | [2] |  |  |
| Covid-19 death rate (20-29) | 1.37 (0.94-1.94) | [9-10] |  |  |
| Covid-19 death rate (30-49) | 10.2 (9.4-11.1) | [9-10] |  |  |
| Covid-19 death rate (50-59) | 55.7 (53.2-58.1) | [9-10] |  |  |
| CVST incidence rate (20-29) | 0.64 (0.29-1.42) | [3] |  |  |
| CVST incidence rate (30-49) | 1.81 (1.29-2.32) | [3] |  |  |
| CVST incidence rate (50-59) | 1.00 (0.62-1.61) | [3] |  |  |
| DIC incidence rate (20-29) | 0.60 (0.34-1.06) | [3] |  |  |
| DIC incidence rate (30-49) | 1.08 (0.26-1.90) | [3] |  |  |
| DIC incidence rate (50-59) | 3.07 (2.50-3.77) | [3] |  |  |
| TEE incidence rate (20-29) | 40.14 (33.75-47.46) | [3] |  |  |
| TEE incidence rate (30-49) | 85.08 (79.40-91.12) | [3] |  |  |
| TEE incidence rate (50-59) | 200.73 (189.54-212.56) | [3] |  |  |
| RR CVST in vaccine users (20-29) | 21.8 (0.28-121.3) | [3] | 44.92 (18.05-92.55) | [3,12-13] |
| RR CVST in vaccine users (30-49) | 3.67 (1.9-6.4) | [3] | 7.98 (5.56-11.11) | [3,12-13] |
| RR CVST in vaccine users (50-59) | 1.40 (0.2-5.1) | [3] | 8.33 (4.31-14.56) | [3,12-13] |
| RR DIC in vaccine users (20-29) | 23.3 (0.30-129.4) | [3] | 6.84 (0.17-38.1) | [3,12-13] |
| RR DIC in vaccine users (30-49) | 2.02 (0.54-5.16) | [3] | 4.14 (2.07-7.42) | [3,12-13] |
| RR DIC in vaccine users (50-59) | 0.23 (0.01-1.27) | [3] | 0.68 (0.14-1.98) | [3,12-13] |
| RR TEE in vaccine users (20-29) | 3.82 (1.91-6.84) | [3] |  |  |
| RR TEE in vaccine users (30-49) | 1.30 (1.03-1.62) | [3] |  |  |
| RR TEE in vaccine users (50-59) | 0.41 (0.29-0.56) | [3] |  |  |
| DIC fatality rate | 0.57 (0.18-0.90) | [3] | 0.53 (0.26-0.79) | [3,12-13] |
| CVST fatality rate | 0.33 (0.13-0.59) | [3] | 0.28 (0.16-0.42) | [3,12-13] |
| TEE fatality rate | 0.22 (0.17-0.29) | [3] |  |  |

DIC: Disseminated Intravascular Coagulation; CVST: Cerebral Venous Sinus Thrombosis; TEE: Thrombo-Embollic Events

**Table 2.** Estimated risk-benefit [B-R] ratio of ChAdOx1 nCoV-19 vaccination.

| Covid-19 deaths prevented per excess death for embolic and thrombotic events |  | Covid-19 deaths prevented per excess death for Disseminated Intravascular Coagulation and Cerebral Venous Sinus Thrombosis |  |  |  |
| --- | --- | --- | --- | --- | --- |
| Based on original estimates |  | Based on original estimates |  | Based on updated estimates |  |
| Age | B-R Ratio (95% UI) | Age | B-R Ratio (95% UI) | Age | B-R Ratio (95% UI) |
| 20-29 | 0.70 (0.27-2.11) | 20-29 | 0.93 (0.06-14.3) | 18-24 | 1.48 (0.33-5.54) |
| 30-49 | 22.9 (10.1-186.4) | 30-49 | 52.2 (16.6-179.1) | 25-49 | 22.7 (10.9-44.7) |
| 50-59 | 1577.1 (1176.9-2121.5) | 50-59 | 3506.7 (166.0-54427.9) | 50-59 | 305 (109.7-851.9) |

**Table 4.** Estimated benefit-risk [B-R] ratio of ChAdOx1 nCoV-19 vaccination, assuming different extents of under-reporting of embolic and thrombotic events.

|  | 30% under-reporting | 50% under-reporting | 80% under-reporting |
| --- | --- | --- | --- |
| Age | B-R Ratio (95% UI) | B-R Ratio (95% UI) | B-R Ratio (95% UI) |
| 18-24 | 0.49 (0.19-1.46) | 0.34 (0.13-1.04) | 0.14 (0.05-0.42) |
| 25-49 | 16.1 (7.10-131.6) | 11.4 (5.03-98.1) | 4.59 (2.03-36.5) |
| 50-59 | 1104.4 (823.2-1483.4) | 788.9 (588.7-1058.7) | 315.6 (235.7-422.6) |

A secondary analysis accounting for the under-reporting of cases shows that even assuming that a substantial proportion of cases was missed by Eudravigilance, the benefit-risk ratio would remain positive for subjects above age 30 (table 3).

Plots of the probabilistic sensitivity analysis clearly show how, in the 30-49 and 50-59 age groups in almost all simulations the number of prevented Covid-19 deaths exceeded that of vaccine-related deaths (Figures 1-6).

**Figure 1.**
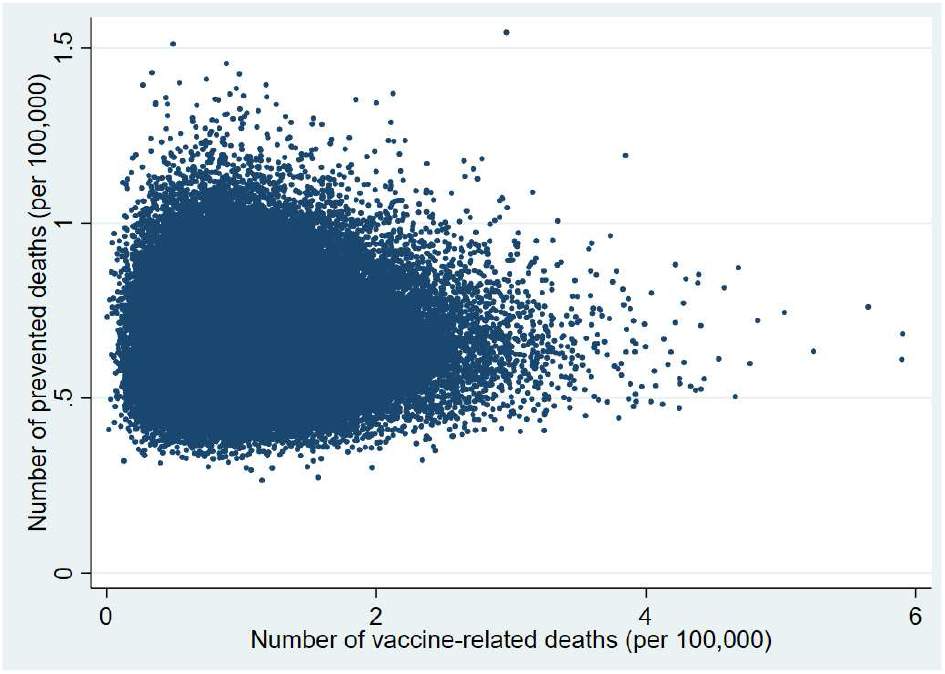
Probabilistic sensitivity analysis. Number of vaccine-prevented deaths for covid-19 (Benefit) vs vaccine-related deaths for Thrombo-Embolic Events (Risk) after 100,000 simulations. Individuals aged 20-29 years, Italy.

**Figure 2.**
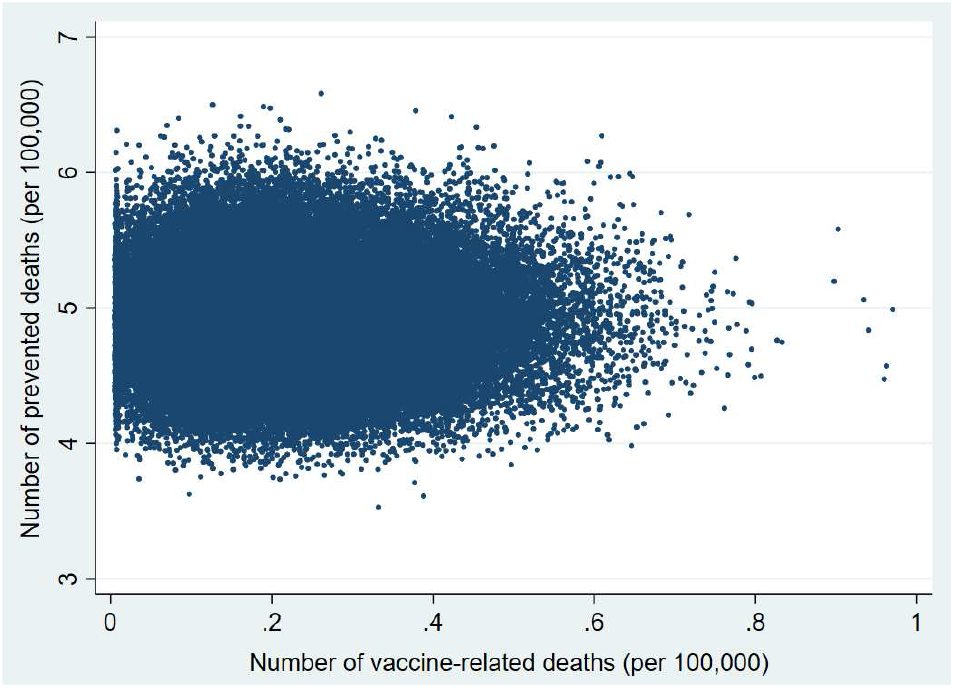
Probabilistic sensitivity analysis. Number of vaccine-prevented deaths for covid-19 (Benefit) vs vaccine-related deaths for Thrombo-Embolic Events (Risk) after 100,000 simulations. Individuals aged 30-49 years, Italy.

**Figure 3.**
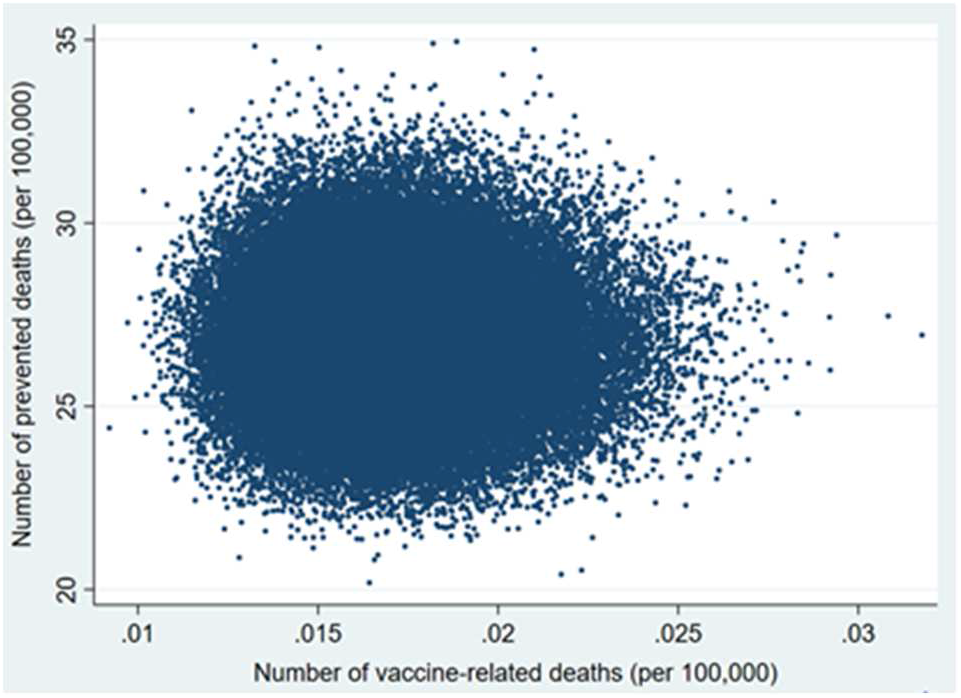
Probabilistic sensitivity analysis. Number of vaccine-prevented deaths for covid-19 (Benefit) vs vaccine-related deaths for Thrombo-Embolic Events (Risk) after 100,000 simulations. Individuals aged 50-59 years, Italy.

**Figure 4.**
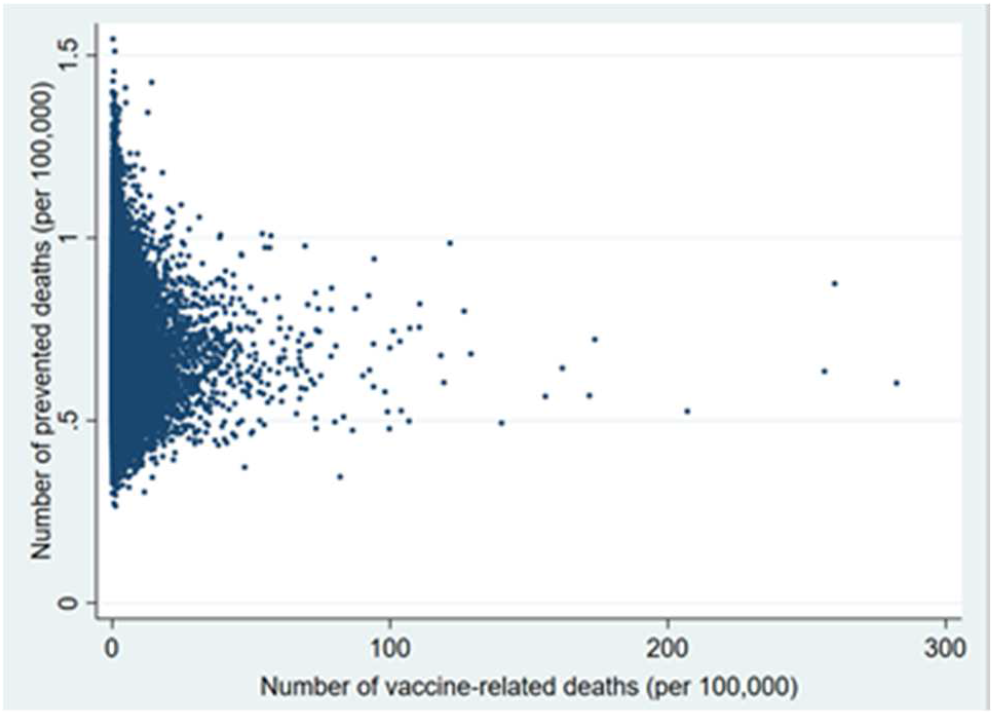
Probabilistic sensitivity analysis. Number of vaccine-prevented deaths for covid-19 (Benefit) vs vaccine-related deaths for Disseminated Intravascular Coagulation and Cerebral Venous Sinus Thrombosis (Risk) after 100,000 simulations. Individuals aged 20-29 years, Italy.

**Figure 5.**
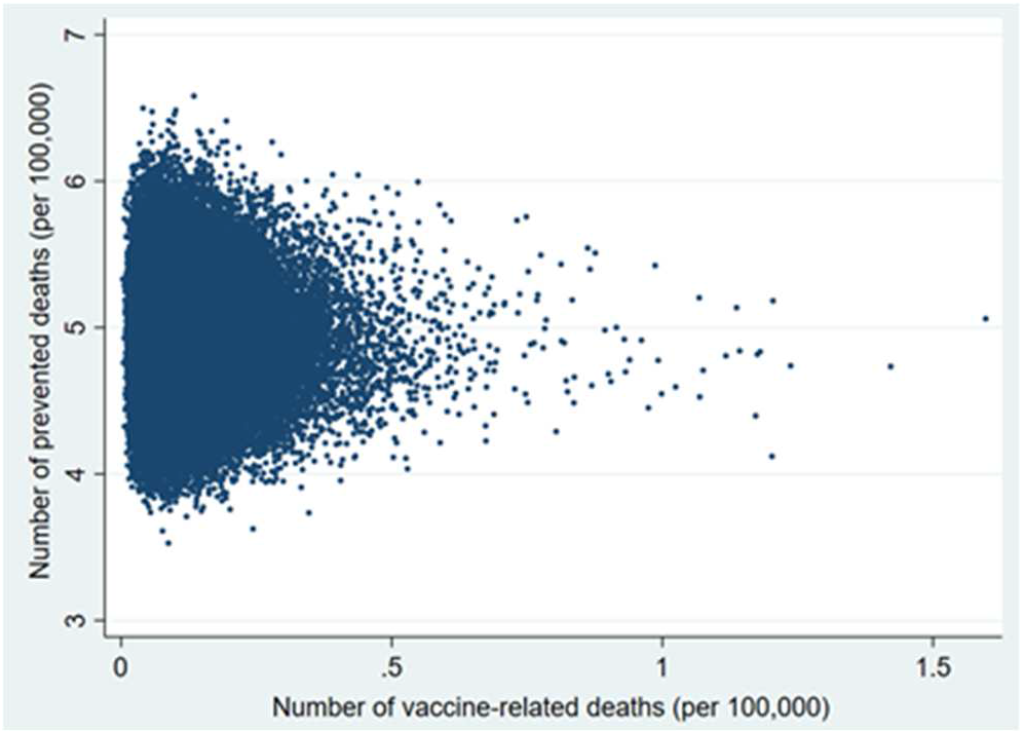
Probabilistic sensitivity analysis. Number of vaccine-prevented deaths for covid-19 (Benefit) vs vaccine-related deaths for Disseminated Intravascular Coagulation and Cerebral Venous Sinus Thrombosis (Risk) after 100,000 simulations. Individuals aged 30-49 years, Italy.

**Figure 6.**
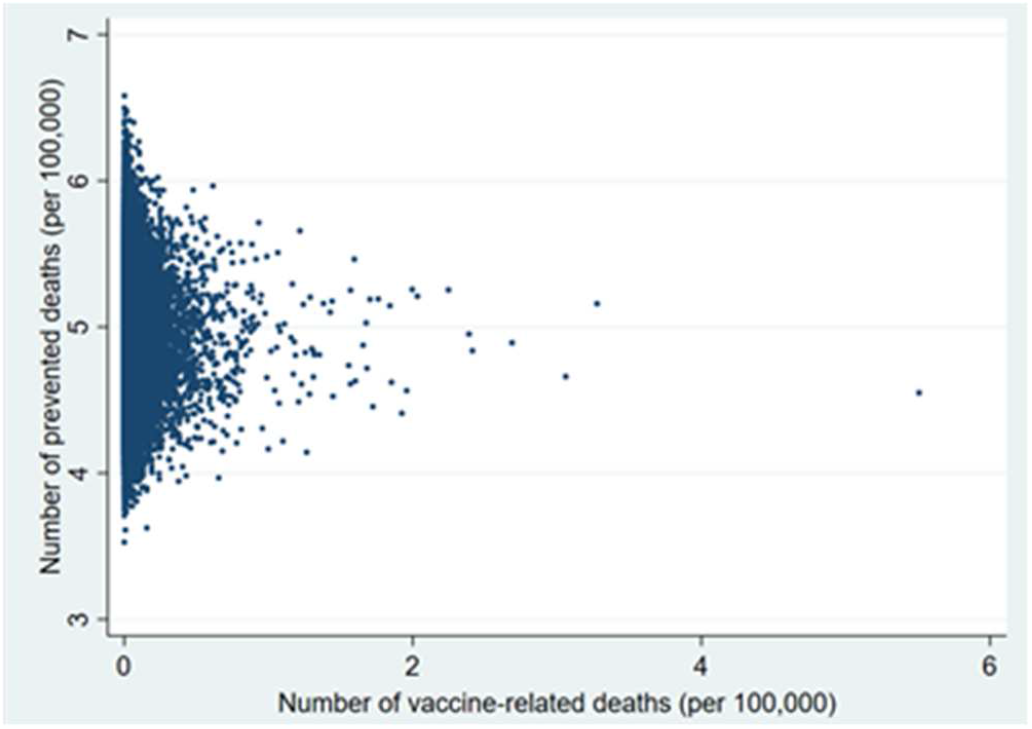
Probabilistic sensitivity analysis. Number of vaccine-prevented deaths for covid-19 (Benefit) vs vaccine-related deaths for Disseminated Intravascular Coagulation and Cerebral Venous Sinus Thrombosis (Risk) after 100,000 simulations. Individuals aged 50-59 years, Italy.

## Discussion

Although Italy, as many other European countries, has limited the use of the ChAdOx1 nCoV-19 vaccine to individuals older than 60 years, the benefits of using this vaccine clearly outweigh the risks for everybody older than 30 years.

The country is currently considering extending again the ChAdOx1 nCoV-19 vaccine to those aged 50-59 years. However, the decision to restrict its use to certain age-groups without a clear directive from EMA has already impacted the immunization campaign greatly, considering that not only the decision itself but also the subsequent media coverage might have profoundly eroded public confidence and increased the risk of vaccine hesitancy [15].

Italy, like many other European countries, due to a mix of factors including EMA delay in granting marketing authorization for all COVID-19 vaccines, shortage in vaccine supply, and logistic problems is currently struggling with the national immunization campaign whilst experiencing the end of a third, very long COVID-19 wave. On the other hand, countries with high vaccination coverage such as the US, the UK, and Israel have seen a steady decline in COVID-19 cases regardless of the type of COVID-19 vaccine predominantly used for the immunization campaign [16].

Although other COVID-19 vaccines have shown a higher efficacy in preventing COVID-19 than the ChAdOx1 vaccine and might be preferred by many of the countries in the long term, when vaccine supply won’t be a problem anymore, there are important factors to consider. Like other vaccines, the ChAdOx1 nCoV-19 vaccine efficacy in preventing COVID-19 complication and deaths is reported to be over 90% [17]. Furthermore, its low market price and its relatively high temperature for storage and transport (2-8 °C), can facilitate immunization even in settings with limited resources.

Our work has some limitations, the main one being represented by the possible under-reporting of the data reported in the Eudravigilance database. As the adverse effects are not always reported, the real cases of thrombotic events may be higher than those detected. As shown in Table 3, however, even in the presence of substantial under-reporting, the results suggest a clear benefit in everybody aged at least 30 years. Another important limitation is related to the fact that we did not produce gender-specific estimates. Future research should evaluate whether this vaccine is associated with larger TEE risk among young women, also considering that some of them take estrogen-progestin contraceptives or hormone replacement therapy.

COVID-19 is having a dramatic impact on the Italian and other European Health Systems. Therefore, the use of ChAdOx1 nCoV-19 vaccine, extending its recommendation to younger population as well, should be a strategic and fundamental part of the immunization campaign considering its safety and efficacy in preventing COVID-19 and its complications.

## Data Availability

The study was based on publicly available aggregate data.

